# The retrospective observational study of clinical outcomes of single dose infusion of warm blood cardioplegia in patients undergoing cardiac surgery

**DOI:** 10.1101/2021.07.15.21260424

**Authors:** Igor I. Chernov, Iliya A. Ivashchenko, Irina A. Mandel

## Abstract

**Objective:** The aim of the study was to assess the safety and efficacy of a normothermic cardioplegia solution N™ use and obtain additional information about dosing regimens during normothermic or mild hypothermic cardiac surgery.

**Methods:** A retrospective observational study included 150 cardio surgery patients. The primary endpoint was the intraoperative acute heart failure development. The secondary endpoints were the postoperative Troponin T concentrations, the need for catecholamine support, and the repeated infusion of a cardioplegia solution.

**Results:** The duration of aortic cross-clamping varied from 17 to 154 minutes, median 59 [interquartile range, 46 - 73] minutes. Spontaneous sinus rhythm recovery was observed in 136 (90.7%) patients. Intraoperative acute heart failure was observed in 1 case. The Troponin T concentrations were 0.331±0.143 ng/mL after surgery. Mortality was 2% (3 patients). Eight patients received an additional volume of N™ solution to maintain asystole. Among 16 patients with a cross-clamp duration greater than 90 minutes epinephrine was used in 3 (18.8%) patients in dose of more than 0.05 mcg/kg/min. Among 134 patients cross-clamp duration less than 90 minutes the catecholamine support was used in 4 (3%) patients, p=0.027.

**Conclusions:** A primary single-dose infusion of cardioplegia solution N™ provides myocardial protection for 59 [interquartile range, 46 - 73] minutes and up to 154 minutes. The catecholamine support in the group of aortic cross-clamp duration less than 90 minutes was used lesser than in the group of aortic cross-clamp duration greater than 90 minutes (3% and 18.8%, respectively). The cardioprotection during cardiopulmonary bypass surgery especially in elderly patients with concomitant disease needs to be confirmed in a future investigations.

## 1 INTRODUCTION

The main goal in cardiac surgery since the 1980s was the reduction of the number of intraoperative and postoperative complications. Aortic clamping during cardiopulmonary bypass (CPB) may lead to ischemia–reperfusion injury which is the main cause of complications and mortality in cardiac surgery. It is understood that myocardial failure due to inadequate protection may occur despite surgical success. Perioperative myocardial infarction is one of the most important adverse events occurring during cardiac surgery.^1-6^ During myocardial ischemia, cardioplegia is the preferred method of myocardial protection. However, decades after its implementation, there is still no consensus on the optimal redosing interval. Shorter re-dosing (15-30 min) has been preferred to longer intervals (45-60 min), but the choice of one approach over another relies more on the surgeon’s preference than on scientific reports.^7^

In 2016 the normothermic cardioplegic blood composition N™, which is explored in this article, was approved for medical use. Recently, a clinical trial was conducted to compare Normacor (N™) and Custodiol (Histidine-tryptophan-ketoglutarate). Similar safety and tolerability were demonstrated of both solutions.^8^

The mechanism of action of the N™ solution is based on the composition of electrolytes, the main of which is potassium. An influx of potassium depolarizes the myocardial membrane causing contraction and thus release and subsequent sequestration of calcium ions resulting in a diastolic arrest. The persistence of potassium reduces the membrane potential and does not allow for adequate repolarization. As the solution diffuses and there is a washout of its components along with products of cellular metabolism, electrical activity begins to appear, and re-dosing of cardioplegia is required if clinically indicated. Potassium, however, is not the only ion in cardioplegia. Other ions such as magnesium, and mannitol all participate in reducing contractility and preserving the myocardium.^9^ N™ protects cardiomyocytes from interstitial edema with mannitol and prevents cellular acidosis with an alkaline pH of the solution. The combination of these factors provides prevention of cellular metabolism disorders.

The upper limit of the duration of aortic clamping and the duration of myocardial protection has not yet been determined due to the limited duration of the operation. To date more than 5000 operations using N™ have been performed in Russia.

The aim of the study was to assess the safety and potential risk of intraoperative acute heart failure (AHF) developing, analyze the efficacy of N™ use and obtain additional information about dosing regimens.

## 2 METHODS

### 2.1 Participants and procedure

This retrospective observational study included all consecutive patients (n=150, 106 men; 58±11.7 years old) undergoing various cardiac procedures using N™ cardioplegic mixture with normothermic (36-37°C) or mild hypothermic (31-35°C) CPB between May 17, 2017 and February 16, 2018. The study was carried out in accordance with The Code of Ethics of the World Medical Association (Declaration of Helsinki) for experiments involving humans. The study was approved by the Institutional Ethical Committee. The Institutional Ethical Committee waived the need for written informed consent to perform this retrospective analysis.

The study included all patients, regardless of the severity of heart disease, various concomitant diseases, overweight and the complexity of surgical intervention. The study protocol did not contain exclusion criteria.

The primary endpoint was the intraoperative AHF development, which was identified as a reduced left ventricle ejection fraction (LVEF) <40% after weaning from the CPB and heart rhythm restoration. The secondary endpoints were the postoperative Troponin T concentrations (6 hours after surgery), the need for catecholamine support and repeated infusion of a cardioplegia solution.

Electrocardiography (ECG) and echocardiography were monitored the day before surgery, during surgery, and 6 hours after surgery.

The anesthetic technique consisted of oral premedication (20 mg phenazepam), induction with 2 mg/kg propofol followed by total intravenous anesthesia with propofol (3 mg/kg/ h). The fentanyl 3 mcg/kg/h (continuous intravenous infusion) was used for analgesia. Neuromuscular blockade was achieved by administering pancuronium bromide (0.15 mg/kg) or vecuronium (0.15 mg/kg). Intravenous heparin (300 IU/kg) was administered immediately before cannulation for CPB and additional doses were given to maintain an activated clotting time of 480 s or greater. Mechanical ventilation was carried out using Primus (Drager, Germany), at a tidal volume of 6 ml/kg with 5 cm H2O positive end-expiratory pressure, and at a respiratory rate of 12/min. The radial artery was cannulated with catheter Arteriofix 20G (BBraun, Germany) for continuous blood pressure monitoring (Siemens-7000, Germany). A temperature sensor was placed into the oesophagus. The ECG (the ST segment abnormalities, arrhythmia episodes), and pulsoxymetry were analyzed with a multifunctional monitor (Siemens-7000, Germany). Echocardiography (Esaote MyLab, Italy) was performed during surgery to monitor the left and right ventricular ejection fraction. Blood analysis of arterial and venous blood for electrolyte balance, acid-base balance and gas balance were performed on Stat Profile CCX (Nova Biomedical, USA) at the beginning and every 30 min of the operation.

### 2.2 Cardioplegia Protocol

N™ cardioplegic solution was mixed with patient’s oxygenated blood. Normothermic cardioplegia was performed by antegrade method through aortic root or coronary arteries separately, or by retrograde method through coronary sinus depending on a type of pathology.

N™ includes two solutions: Solution No.1 (high-potassium concentration) mixed with normothermic oxygenated blood for achievement of asystole and Solution No.2 (low-potassium) mixed with normothermic oxygenated blood for maintain asystole.

Composition per 1000 ml of solution:

Solution No. 1. Active substances: Potassium chloride: 7.450 g (potassium ions: 3.90 g), Magnesium sulphate heptahydrate: 2.340 g (magnesium ions: 0.23 g), Mannitol: 35.900 g.

Additive: Trometamol (trishydroxymethylaminomethane): 0.500 g, hydrochloric acid: up to pH 7.6-8.0, water for injection: up to 1000 ml. The theoretical osmolarity of the solution is 439.95 mosm/L.

Solution No. 2. Active substances: Potassium chloride: 2.125 g, Magnesium sulphate heptahydrate: 2.340 g, Mannitol: 58.280 g.

Additive: Trometamol (trishydroxymethylaminomethane): 0.500 g, hydrochloric acid: up to pH 7.6-8.0, water for injection: up to 1000 ml. The theoretical osmolarity of the solution is 414.94 mosm/L.

The cardioplegic solution was delivered to the aortic root using a roller pump and a tube system as part of CPB (Figure 1).

**Figure 1.**
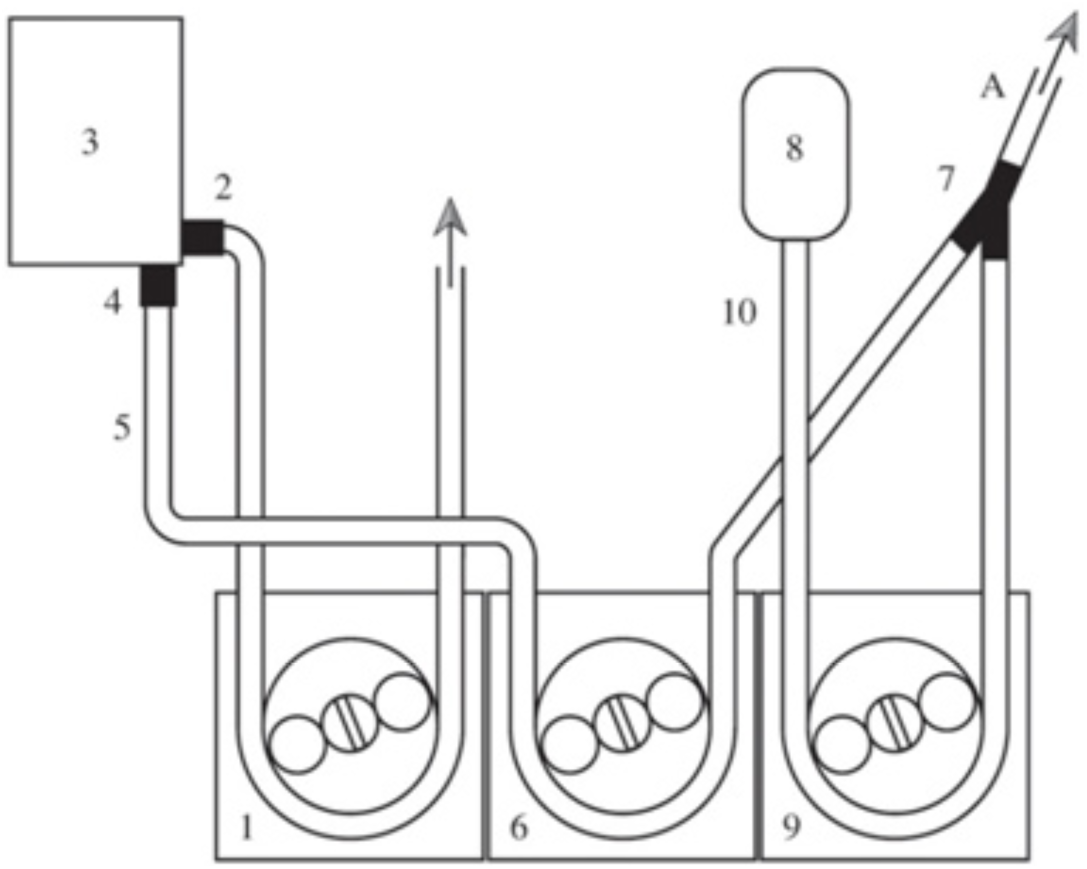
Scheme of the administration of N™ (Solution No. 1 or Solution No. 2), and the oxygenated blood cardioplegic mixture 1 - arterial pump; 2 - connector for arterial perfusion, 3 - oxygenator; 4 - connector for coronary perfusion; 5 - coronary perfusion line; 6, 9 - roller pumps for cardioplegic solution to the heart–lung machine; 7 – T-joint connecting the blood and N™ solution; 8 - vial with Solution No. 1 or with Solution No. 2; 10 - the main line for coronary perfusion with N™ solution; A – a common cardioplegic line.

The cardioplegic mixture completely entered into the cardiotomic reservoir. The main criterion for the correct dose of N™ and the indicator of the cardioplegic efficacy was the time until the onset of asystole. In case of antegrade perfusion if the heart has continued to beat after 1 or 2 minutes from the cardioplegia beginning the surgeon checked the aorta cross-clamp and the aortic valve insufficiency. If the solution does not enter the coronary bed in sufficient volume we switched to a retrograde perfusion method. Antegrade perfusion of the cardioplegic mixture of N™ and oxygenated blood was performed in 1:2 ratio. The hematocrit was 23±3. The infusion rate of the mixture according to the antegrade method was 300 ml/min. A total volume of 400 ml of N ™ solution was injected.

Retrograde perfusion of the cardioplegic mixture of N™ and oxygenated blood was performed in 1:2 ratio. The infusion rate of the mixture through the coronary sinus after applying a cross-clamp to the aorta was 100-150 ml/min (pressure didn’t exceed 50 mm Hg).

If ventricular activity appeared at the main stage of the operation we maintained asystole by perfusion of the N ™ cardioplegic mixture with oxygenated blood in a ratio of 1:2 using one of the following methods:

- Antegrade method with the infusion rate of N™ cardioplegic mixture 150 ml/min until asystole was achieved;
- Retrograde method at the infusion rate of N™ cardioplegic mixture of 100-150 ml/ min until the asystole was achieved.

The appearance of atrial activity during the main stage of the operation did not affect the efficacy of myocardial protection.

The effect of cardioplegia was finished immediately after the cross-clamping of aorta was removed. In this case the restoration of cardiac activity occurred mainly through brady-cardia due to residual manifestations of hyperkalemia. Bradycardia was stopped as soon as the concentration of potassium ions normalized.

Normothermic surgical procedures in 143 (95.3%) patients (36-37°C) and mild hypothermic in 7 (4.7%) patients (31-35°C) were performed. Antegrade method of cardioplegic mixture delivery was used in 146 (97.3%) patients. Consecutively antegrade and retrograde methods were used in 4 (2.7%) patients.

Initially N™ mixture was administered in all cases. The mean value of the primary volume of Solution No. 1 was 415.33 ± 68.28 ml. Repeated infusion of solution No. 1 was used in 7 cases of premature restoration of ventricular activity in order to maintain asystole in a volume of 200-400 ml. Solution No. 2 was used in one case (100 ml) to maintain asystole when ventricular activity was restored prematurely.

### 2.3 Statistical analysis

Data processing and comparison of variables have been performed by appropriate statistical methods. Comparison by qualitative characteristics was performed by the Fischer’s exact test, difference between groups was considered significant if p-value was <0.05. Continuous data were checked for normality with D’Agostino-Pearson omnibus normality test. Comparison between groups was performed using Unpaired T test in case of normal distribution and Mann-Whitney test otherwise. Data analyses were performed using GraphPad Prism 5 (GraphPad Software Inc., CA, USA).

## 3 RESULTS

### 3.1 Patients population

The group of 150 adult patients (60 [52.75 - 66.25] years old, male 84 (56%)) was observed. The youngest patient was 22 years old and the oldest one was 80 years old. Body mass index (BMI) was 27.7 [24.9 - 33.7] kg/m^2^, varied from 18 to 43 kg/m^2^. According to the European System for Cardiac Operative Risk Evaluation (Euro-SCORE)^10^ a predicted mortality rate was 3.29 [1.79 - 5.90]%.

### 3.2 Preoperative Diagnosis

The study included 2 (1.3%) patients with a very low ejection fraction of the left ventricle (lesser than 30% according to Simpson J.S.); 26 (17.3%) patients who had myocardial infarction at least 6 months before surgery; 6 (9%) patients had post infarction left ventricular aneurysm. The baseline characteristics of the patients are presented in Table 1 and Figure 2A.

**Table 1.** Baseline Characteristics of patients.

| **Characteristic** | **Overall** |  | **LVEF**<br>**≥ 55%** |  | **LVEF**<br>**45 -55%** |  | **LVEF**<br>**30-45%** |  | **LVEF**<br>**< 30%** |  |
| --- | --- | --- | --- | --- | --- | --- | --- | --- | --- | --- |
|  | N | % | N | % | N | % | N | % | N | % |
| *Anamnesis* |  |  |  |  |  |  |  |  |  |  |
| Myocardial infarction<br>6 month before surgery | 26 | 17.3 | 8 | 5,3 | 9 | 6.0 | 7 | 4.7 | 2 | 1.3 |
| LV PI aneurysm | 9 | 6.0 | 2 | 1,3 | 3 | 2.0 | 3 | 2.0 | 1 | 0.7 |
| NYHA (I; II; III; IV) | 20 | 13.3 | 3; 10;<br>0; 0 |  | 1; 2; 1;<br>0 |  | 2; 0;<br>1; 0 |  | 0; 0;<br>0; 0 |  |
| Circulatory insufficiency (I; IIA; IIB; III) | 115 | 76.7 | 17;<br>43; 7;<br>5 |  | 2; 19;<br>2; 3 |  | 2; 11;<br>2; 0 |  | 0; 1;<br>1; 0 |  |
| Killip class (I; II; III; IV) | 20 | 13.3 | 3; 10;<br>0; 0 |  | 1; 2; 1;<br>0 |  | 2; 0;<br>1; 0 |  | 0; 0;<br>0; 0 |  |
| *Type of diagnosis* |  |  |  |  |  |  |  |  |  |  |
| CAD | 54 | 36.0 | 22 | 14.7 | 19 | 12.7 | 11 | 7.3 | 2 | 1.3 |
| Acquired heart disease | 129 | 86.0 | 85 | 56.7 | 28 | 18.7 | 14 | 9.3 | 2 | 1.3 |
| Congenital heart disease | 17 | 11.3 | 13 | 8.7 | 1 | 0.7 | 3 | 2.0 | 0 | 0.0 |
| Cardiomyopathy | 4 | 2.7 | 1 | 0.7 | 1 | 0.7 | 2 | 1.3 | 0 | 0.0 |
| Pathology of the aorta | 27 | 18.0 | 20 | 13.3 | 5 | 3.3 | 2 | 1.3 | 0 | 0.0 |
| Pathology of other vessels | 15 | 10.0 | 10 | 6.7 | 5 | 3.3 | 0 | 0.0 | 0 | 0.0 |
| Combined pathology | 70 | 46.7 | 36 | 24.0 | 21 | 14.0 | 11 | 7.3 | 2 | 1.3 |

CAD - Coronary artery disease; LVEF – left ventricular ejection fraction according to Simpson J.S.; NYHA - the New York Heart Association; LV PI aneurism - post infarct left ventricular aneurysm

**Figure 2.**
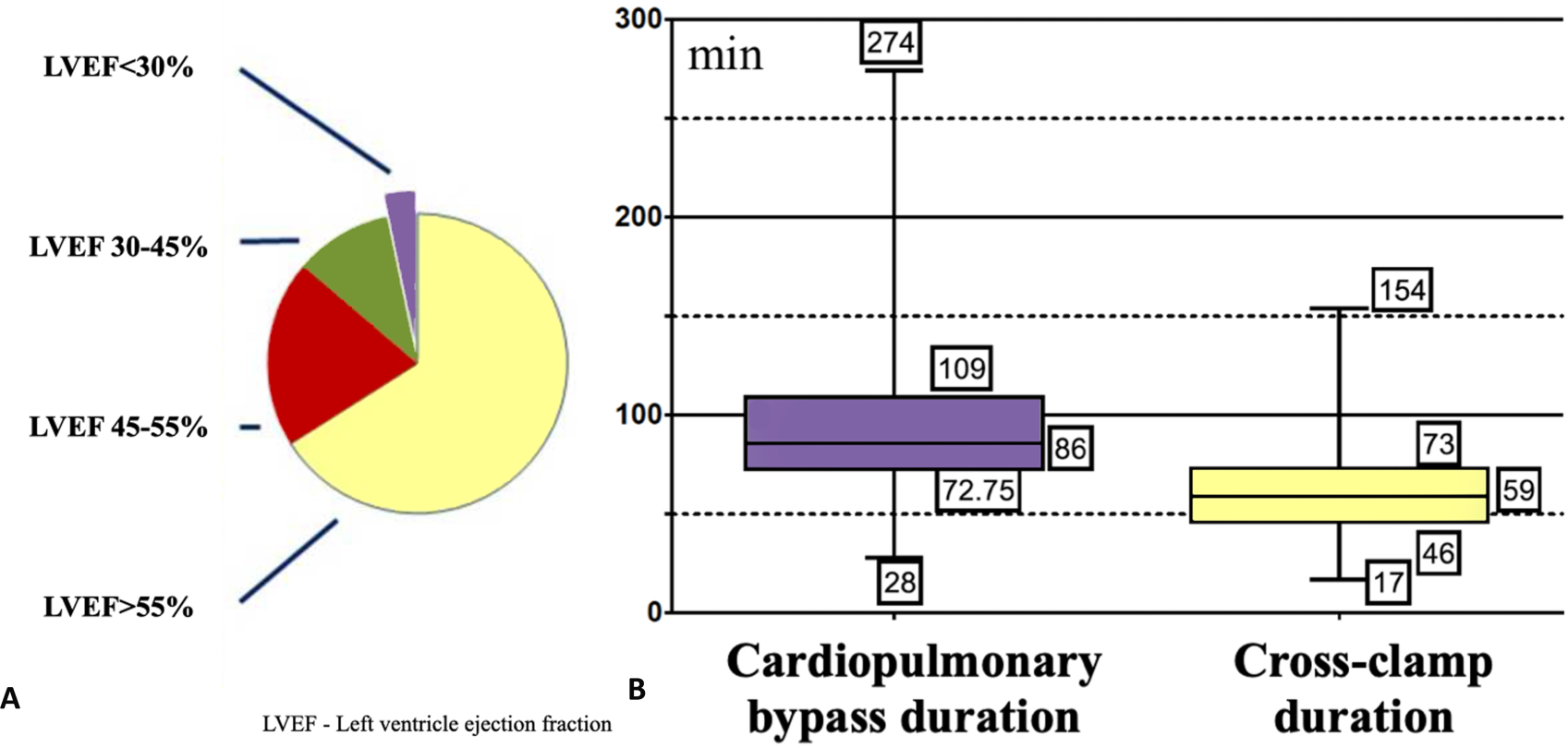
A - Pie diagram of the preoperative status of the patients (n=150); LVEF - left ventricular ejection fraction. B - Box-and-whisker plots of the cardiopulmonary bypass and cross-clamp duration (n=150), minutes. Boxes represent median values with interquartile range. The lower limit of the box represents the 25th percentile. The upper limit of the box represents the 75th percentile. The ends of whiskers represent the minimum and maximum of all of the data.

### 3.3 Type of surgical intervention

The types of surgery are shown in Table 2. Operations for acquired heart defects were performed in 131 (87.3%) patients. Congenital heart disease in 19 (12.7%) patients. Out of them, combined surgical procedures with tricuspid valve repair were performed in 65 (43.3%) patients, and coronary artery bypass graft surgery were performed in 51 (34%) patients. Correction of aorta pathology was performed in 25 (16.7%) cases.

**Table 2.**
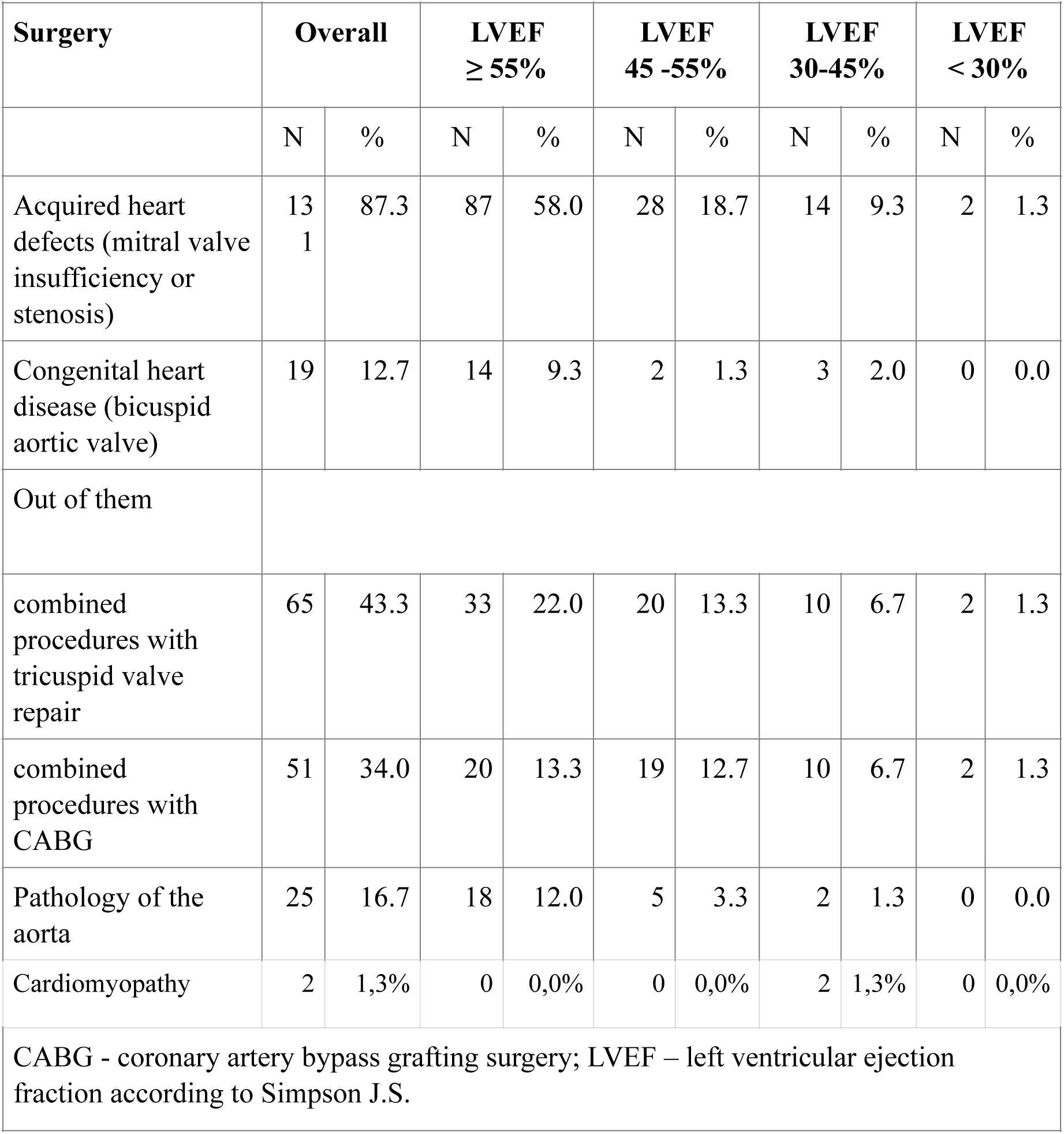
Surgery Type.

### 3.4 Intraoperative period

CPB time in 150 surgical procedures ranged from 28 to 274 minutes (Table 3). In 90 (60%) patients the CPB time was less than 90 minutes. In 60 (40%) cases the CPB time was more than 90 minutes, including 25 (16.7%) cases of more than 120 minutes.

**Table 3.** Surgery characteristics (n=150)

| Surgery characteristics | N | Mean |  | SD | MIN | 25% | MEDIAN | 75% | MAX |
| --- | --- | --- | --- | --- | --- | --- | --- | --- | --- |
| CPB duration, min | 150 | 94.1 | ± | 38.3 | 28 | 72.75 | 86 | 109 | 274 |
| Cross-clamp duration, min | 150 | 61.3 | ± | 24.4 | 17 | 46 | 59 | 73 | 154 |
| Distribution of patients by duration of CPB |  |  |  |  |  |  |  |  |  |
| CPB duration, min |  |  |  | n |  |  |  | % |  |
| ≤90 |  |  |  | 90 |  |  |  | 60.0 |  |
| 91-120 |  |  |  | 35 |  |  |  | 23.3 |  |
| >120 |  |  |  | 25 |  |  |  | 16.7 |  |
| Total |  |  |  | 150 |  |  |  | 100.0 |  |
| Distribution of patients by cross-clamp duration |  |  |  |  |  |  |  |  |  |
| Cross-clamp duration, min |  |  |  | n |  |  |  | % |  |
| ≤60 |  |  |  | 81 |  |  |  | 54.0 |  |
| 61-90 |  |  |  | 53 |  |  |  | 35.3 |  |
| 91-120 |  |  |  | 12 |  |  |  | 8.0 |  |
| 120 |  |  |  | 4 |  |  |  | 2.7 |  |
| Total |  |  |  | 150 |  |  |  | 100.0 |  |
| CPB - cardiopulmonary bypass |  |  |  |  |  |  |  |  |  |

The aortic cross-clamp duration varied from 17 to 154 minutes, Figure 2B. The cardiac ischemia time was lesser then 60 minutes in 81 (54%) patients. Long surgical procedures with a cross-clamp duration of more than 120 minutes were performed in 4 (2.7%) patients. Two patients who had a long period of cross-clamping did not need additional N™ infusion to maintain asystole. Spontaneous sinus rhythm recovery was observed in all 4 cases of long period of ischemia. Catecholamine support with epinephrine at a dose of more than 0.05 µg/ kg/min was used in 2 out of 4 cases.

The vast majority of patients received single dose of 400 ml of N™. A correlation between the volume of the cardioplegia solution and the cross-clamp duration has not been established.

Eight patients received an additional volume of N™ Solution No. 1 (200 ml to 400 ml) at the initial infusion and 8 patients received an additional volume of N™ Solution No. 1 (100 to 400 ml) to maintain asystole during surgery. N™ Solution No. 2 was used once at the dose of 100 ml to maintain asystole during surgery. The use of the additional volume of N™ solution was independent of the cross-clamp duration and the scope of surgical intervention, even in patients with cross-clamp duration of 120 minutes or more. The reason of additional infusion of N™ solution was the incomplete infusion of the entire volume at the first time. This was either due to incomplete aortic cross-clamping or incomplete closure of the aortic valve due to valve defects.

The N™ solution provided cardioprotection for the entire period necessary to complete the surgery. Undesirable reactions in the application of N™ solution were not detected.

### 3.5 Clinical outcomes

The myocardium activity recovery time was 95±11 seconds. In other patients sinus rhythm restored after ventricular fibrillation and defibrillation procedure.

There were no significant difference in Troponin T concentrations after surgery (0.331±0.143 ng/mL) compared with Troponin T concentrations before surgery (0.129±0.038 ng/mL), p=0.092.

In all 150 surgical procedures cardiac arrest type was asystole. Spontaneous sinus rhythm recovery was observed in 136 (90.7%) cases, including 2 patients with LVEF lesser than 30%. Both patients with LVEF lesser than 30% did not require epinephrine support in a dose exceeding 0.05 mcg/kg/min. No arrhythmia was recorded.

Intraoperative AHF occurred in 3 cases. All of them had LVEF more than 55% before surgery. In first two cases multiple organ failure developed after the surgery. In last case a cardiovascular insufficiency developed. This patient received exrtacorporal membrane oxygenation for a ten days and died on 34th day. No cases of stroke, renal, hepatic, pulmonary failure were reported (Table 5).

**Table 4.**
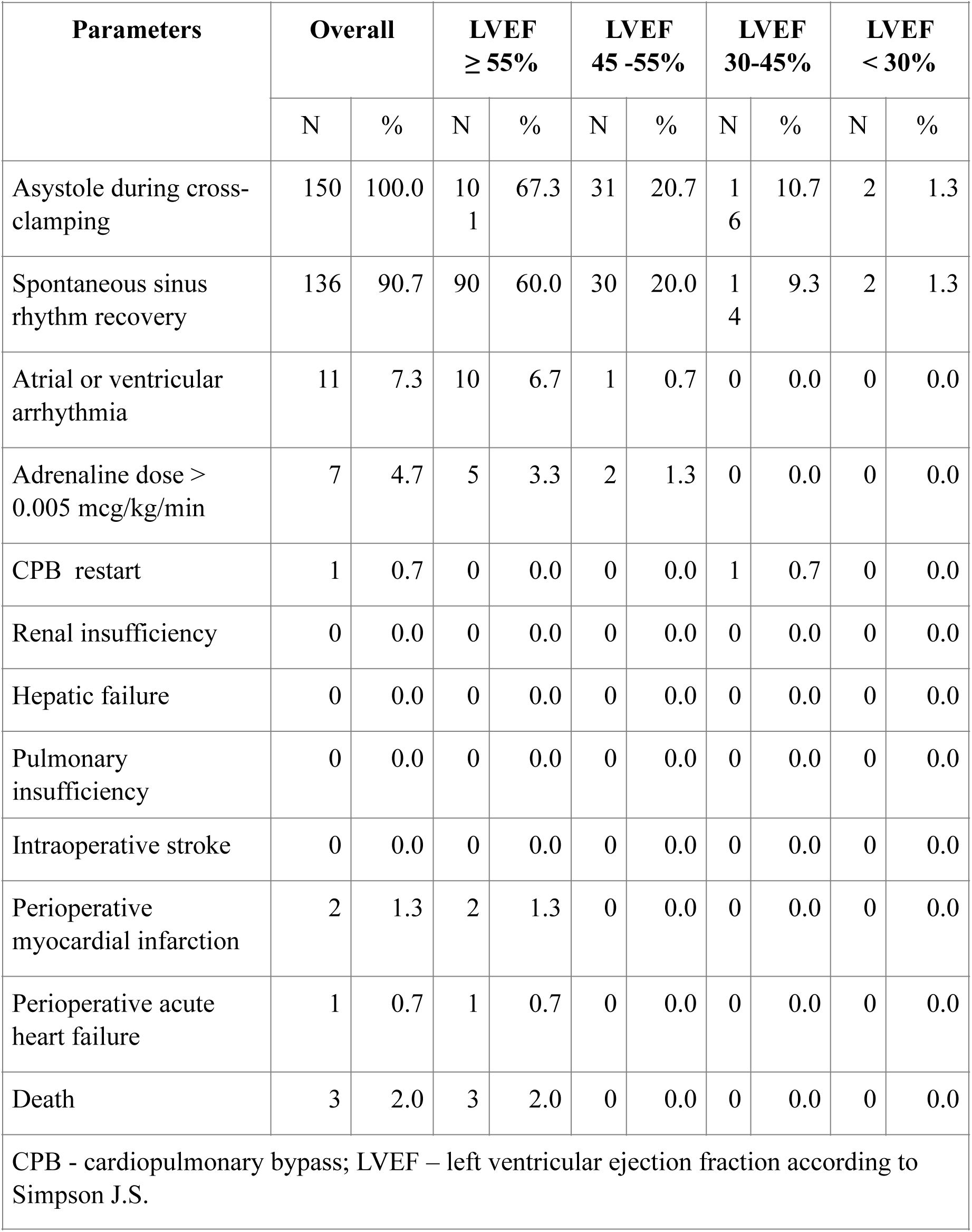
Clinical outcomes.

**Table 5.** Characteristics of postoperative period (n=150)

| Table 5 Characteristics of postoperative period (n=150) |  |  |  |  |  |  |  |  |  |  |
| --- | --- | --- | --- | --- | --- | --- | --- | --- | --- | --- |
| parameters | N | % | Mean |  | SD | MIN | 25% | MEDIA<br>N | 75% | MAX |
| Intensive care unit, hours | 150 | 100 | 43.9 | ± | 78.3 | 15 | 19 | 22 | 42 | 820 |
| Length of hospital stay, days | 150 | 100 | 8.6 | ± | 4.0 | 4 | 6 | 7 | 10 | 34 |
| Distribution of patients by intensive cure unit stay duration |  |  |  |  |  |  |  |  |  |  |
| Intensive care unit, hours |  |  |  |  | n |  |  | % |  |  |
| <24 |  |  |  |  | 99 |  |  | 66.0 |  |  |
| 24-48 |  |  |  |  | 26 |  |  | 17.3 |  |  |
| 48-72 |  |  |  |  | 9 |  |  | 60 |  |  |
| > 72 |  |  |  |  | 16 |  |  | 10.7 |  |  |
| Total |  |  |  |  | 150 |  |  | 100.0 |  |  |
| Distribution of patients by length of hospital stay |  |  |  |  |  |  |  |  |  |  |
| Length of hospital stay, days |  |  |  |  | n |  |  | % |  |  |
| ≤ 7 |  |  |  |  | 78 |  |  | 52.0 |  |  |
| 7-12 |  |  |  |  | 55 |  |  | 36.7 |  |  |
| > 12 |  |  |  |  | 17 |  |  | 11.3 |  |  |
| Total |  |  |  |  | 150 |  |  | 100.0 |  |  |

Defibrillation was performed in 14 (9.3%) patients after aorta declamping. Arrhythmia was recorded in 11 cases (7.3%), 10 of them had LVEF more than 55%. Intraoperative catecholamine support with the epinephrine at a dose of more than 0.05 mcg/kg/min was required in 7 (4.7%) patients (all of them had the LVEF more than 45%).

A significant difference was observed in the distribution of patients according to the catecholamine support (epinephrine more than 0.05 mcg/kg/min) in groups of cross-clamp duration < 90 minutes and > 90 minutes (p=0.027). Among 16 patients with cross-clamp duration > 90 minutes the catecholamine support (epinephrine more than 0.05 mcg/kg/min) was used in 3 (18.8%) cases. Among 134 patients with cross-clamp duration < 90 min the catecholamine support was used in 4 (3.0%) cases. The frequency of administration of epinephrine in doses of more than 0.05 mcg/kg/min in both groups indicated their extremely rare use despite the initially severe patients. The length of catecholamine support was 6.1±1.9 hours.

The median ICU length of stay was 22 hours, only 16 (10.7%) patients spent more than 72 hours in the ICU (Table 5). The ICU length of stay did not statistically significant differ in the groups with short and long cross-clamp duration. This indicates effective heart protection with the N™ solution for any duration of cross-clamping (p=0.679). The length of stay in hospital was 7 [6; 10] days. Seventeen patients (11.3%) spent at the hospital more than 12 days.

### 3.6 Laboratory Parameters

Analysis of laboratory data of arterial and venous blood for electrolyte balance, acid-base balance and gas balance indicates physiological changes during the operation, with the exception of the concentration of potassium ions at the main and final stages of the operation (according to the N™ mechanism of action). All of the above mentioned parameters returned to normal as soon as the operation completed (Figures 3-6).

**Figure 3.**
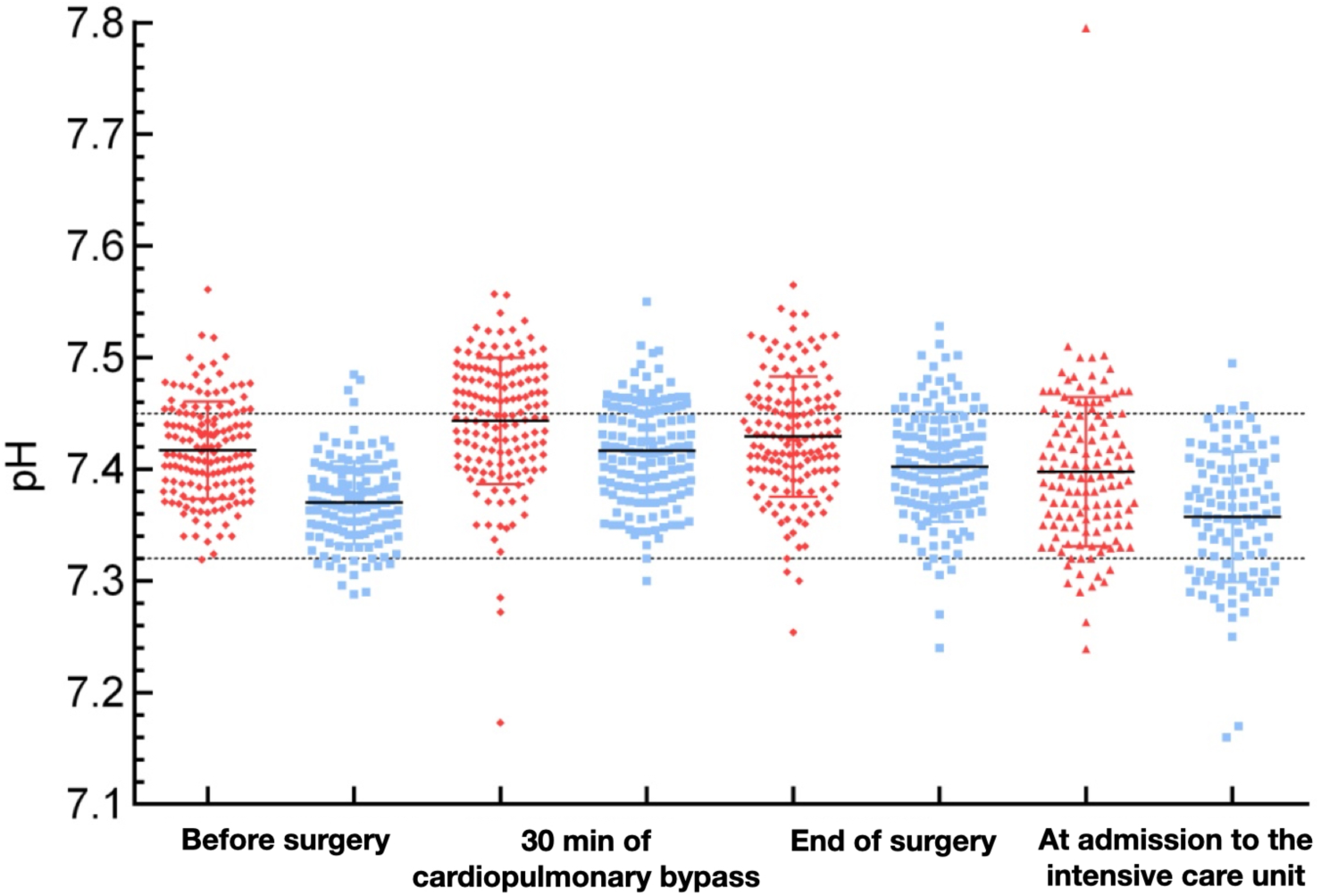
pH levels of arterial (red dots) and venous (blue dots) blood of the patients. Black line represent median values. Dash lines represents the normal range of the parameter. Each dot represents one patient.

**Figure 4.**
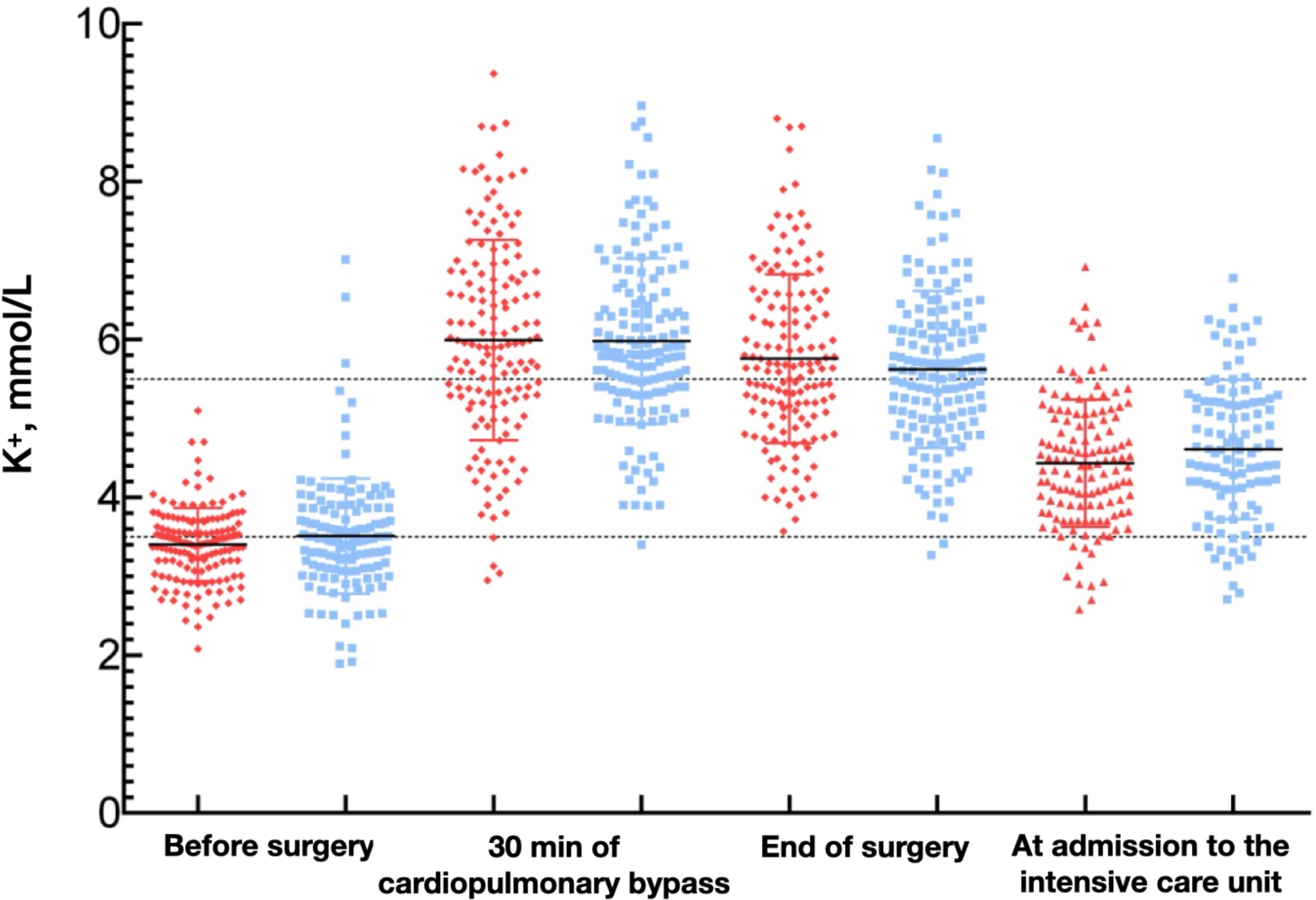
Potassium ions concentrations in arterial (red dots) and venous (blue dots) blood of the patients. Black line represent median values. Dash lines represents the normal range of the parameter. Each dot represents one patient.

**Figure 5.**
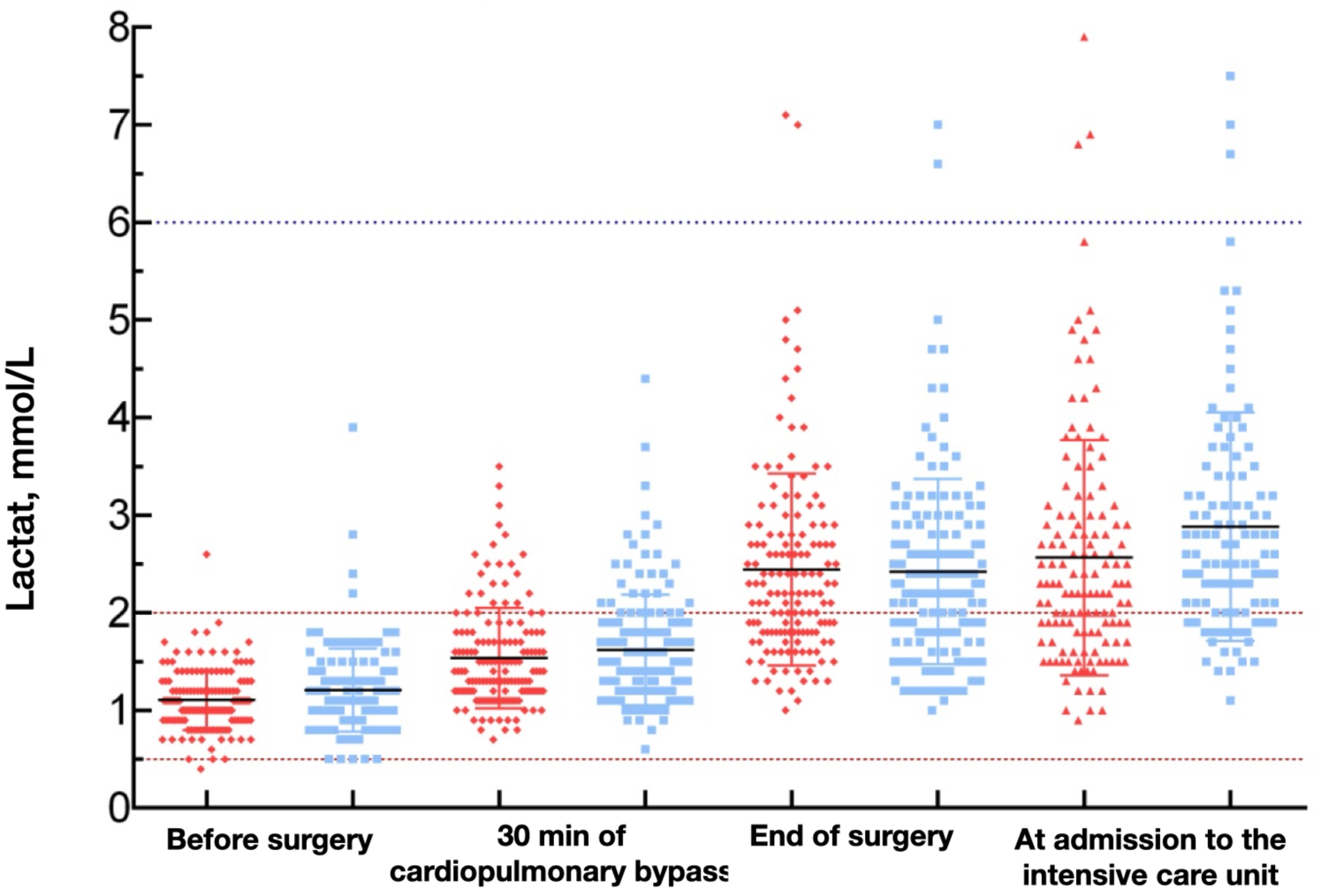
Lactate concentrations in arterial (red dots) and venous (blue dots) blood of the patients. Black line represent median values. Dash lines represents the normal range of the parameter. Each dot represents one patient.

**Figure 6.**
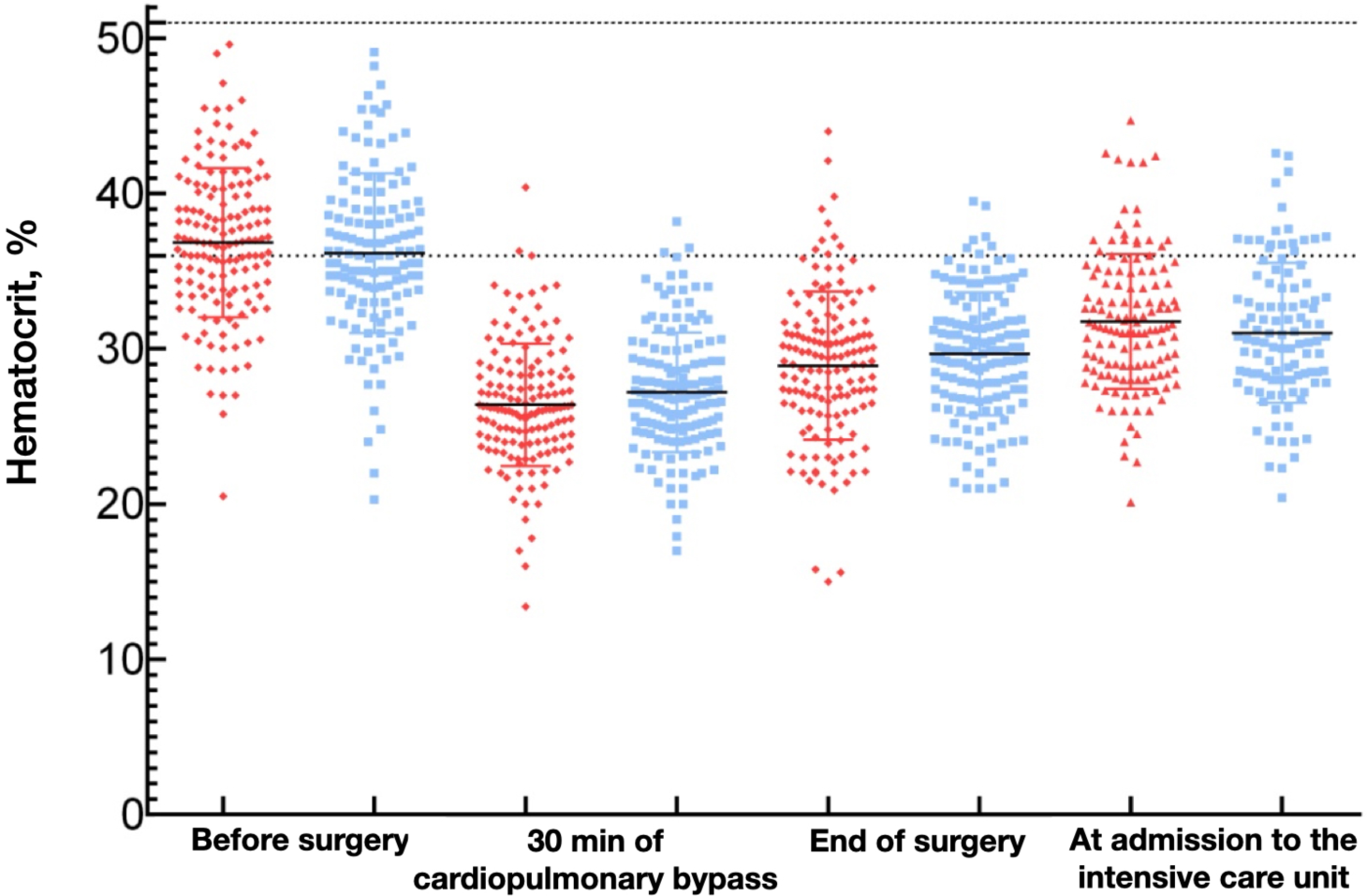
Hematocrit in arterial (red dots) and venous (blue dots) blood of the patients. Black line represent median values. Dash lines represents the normal range of the parameter. Each dot represents one patient.

### 3.7 Clinical case

As a clinical case we present one patient with a cross-clamp duration of 154 minutes, where additional infusion of the N™ solution was not required. Patient P. underwent the following surgery: replacement of the ascending aorta and aortic valve. The 400 ml of the cardioplegic solution was used to protect the heart in a ratio of 1 part of cardioplegic solution and 2 parts of the oxygenated blood. The duration of infusion was 5 minutes. Cardiac arrest occurred at a first minute. Sinus rhythm recovered within four minutes after the aorta cross-clamp was removed. Ventricular activity was not restored during the entire period of cross-clamping. Subsequent infusion of N™ to maintain asystole was not required. The patient was weaned from the CPB with the first attempt. Arrhythmia was not noted. No intraoperative complications were observed. The patient was transferred to the ICU without catecholamine support.

## 4 DISCUSSION

Three (2%) patients developed perioperative AHF. Similar 30-day mortality rates (from 1% to 4%) have been reported in other large cardiac surgery studies.^11-13^

Seven cases (4.7%) of epinephrine administration at the dose of more 0.05 mcg/kg/ min were observed. This indicates the cardioprotective properties of the N™ solution. Analysis of the Del Nido method by Ota T. et al. (2015) indicated that postoperative inotropic support was required for 11 patients (20.4%) in the Del Nido group and for 13 patients (24.1%) in the control group (p=0.82).^14^

Intervals for the infusion of normothermic cardioplegic solutions have been discussed in many publications. Nardi P. et al. (2018) indicated that “warm blood requires a shorter interval of infusion to achieve better protection of myocardium”.^4^ Lichtenstein S.V. et al. (2018) demonstrated that the safe interval for infusion of normothermic blood cardioplegia was 13 minutes.^15^ Ghazy T. et al. (2009) indicated that “the standard in the literature is still the repeated infusion of cardioplegic blood solution every 15 minutes”.^16^

Molina GR et al. (2018) noted that once cross-clamp is applied, and infusion of cold blood cardioplegia has begun, typical ECG changes can be observed in one to two minutes time and an isoelectric ECG recording is obtained after 3 to 4 minutes that lasts for at least 90 minutes or longer. Unless aortic clamping failure occurs, repeated doses of cardioplegia are not required. Other aspects of myocardial protection should be observed such as preventing left ventricle overdistension and avoiding rewarming of the heart. Continuous topical cooling with iced saline solution or slush is advisable.^17^ We didn’t use neither topical cooling, no cold solution. Cardioprotection was provided by the composition of the solution.

The aim of this study was the assessment of possibility and safety of the single dose infusion of normothermic blood cardioplegia in cardiac surgery. The observational study showed that cardioplegia with a single dose infusion of N™ solution does not increase the development of postoperative myocardial infarction. This may indicate that protection of the myocardium with the N™ solution may be sufficient, double infusion may not be required in most procedures. Further studies are needed to establish the safe limit of action of single dose of normothermic blood cardioplegia.

Optimal dosing and timing of del Nido cardioplegia has not been definitively established for adult patients. Typically, only one dose of cardioplegia is given over the entire ischemic period assuming its duration is limited to approximately 90 minutes or less, although this time cutoff has not been tested experimentally and requires further investigation.^18^ Our study assessed standard clinical practice using N™ solution and obtained additional information. One hundred fifty operations were performed under normothermic blood cardioplegia with a single infusion of 400 ml of N™ solution which provided myocardial protection for 59 [46 - 73] minutes and up to 154 minutes of aortic cross-clamping. In 7 cases ventricular activity was restored and infusion of the solution No. 1 was repeated to maintain asystole. Solution No. 2 was used in 1 case. This indicates the possibility of a complete rejection of the use of solution No. 2 for cardioplegia. Asystole may be maintained with repeated infusion of Solution No. 1.

Subsequent infusion of the solution was necessary due to incomplete infusion of the entire volume of mixture at the beginning of the main stage of the operation due to incomplete aorta cross-clamping or incomplete aortic valve closure. It was never necessary to infuse a repeated dose of N™ more than once to maintain asystole even if the duration of the cross-clamp exceeds 120 minutes.

Our analysis has several limitations. This is a single center report. The study was neither comparative nor randomized. This study does not elucidate a clear mechanism of action for N™ solution. We does not provide data regarding the N-terminal pro-brain natriuretic peptide as a marker of myocardial injury. The inclusion of patients with comorbidities in separate groups could add a confounding variable in the study. This study is limited in the duration of followup within 1 month. This work represents a pilot clinical study of acute heart failure development and catecholamine support and requires confirmation by large clinical studies to verify the efficacy of cardioplegia based on N™ solution in clinical settings involving ischemia–reperfusion injury.

## 5 CONCLUSIONS

Normothermic blood cardioplegia solution N™ can be used both for standard cardiac surgery and for patients with high risk of intraoperative complications associated with severe cardiovascular insufficiency or severe concomitant diseases. The safety of the first dose might be sufficient for cardioprotection during the whole cross-clamping time. A primary single dose infusion of cardioplegia solution N™ provides myocardial protection for 59 [46; - 73] and up to 154 minutes. The catecholamine support in case of cross-clamp duration time less than 90 minutes has been rarely used. Cardioplegia solution N™ provides support for physiological myocardium metabolism and therefore adequate myocardial protection in the intraoperative period. Further studies are still needed to reveal the safe limit of single dose of cardioplegia solution.

## Data Availability

The datasets analyzed during the current study are available from the corresponding author on reasonable request.

## Glossary

AHF: Acute heart failure
BMI: body mass index
CPB: cardiopulmonary bypass
ICU: intensive cure unit
LVEF: left ventricle ejection fraction

## Acknowledgements

The authors thank «Federal Center for Cardiovascular Surgery» of Astrakhan for assistance with this work.

## REFERENCES

1. Buckberg GD. Antegrade/Retrograde Blood Cardioplegia to Ensure Cardioplegic Distribution: Operative Techniques and Objectives. J Card Surg. 1989; 4(3):216–38.

2. Salerno TA, Houck JP, Barrozo CA, et al. Retrograde Continuous Warm Blood Cardioplegia: A New Concept in Myocardial Protection. Ann Thorac Surg. 1991; 51:245–7.

3. Calafiore AM, Teodori G, Mezzetti A, et al. Intermittent Antegrade Warm Blood Cardioplegia. Ann Thorac Surg. 1995; 59:398–402.

4. Nardi P, Pisano C, Bertoldo F, et al. Warm blood cardioplegia versus cold crystalloid cardioplegia for myocardial protection during coronary artery bypass grafting surgery. Cell Death Discov. 2018; 4:23.

5. Durandy YD, Hulin SH. Normothermic Bypass in Pediatric Surgery: Technical Aspect and Clinical Experience with 1400 cases. ASAIO J 2006; 52(5):539–42.

6. Martin TD, Craver JM, Gott JP, et al. Prospective, Randomized Trial of Retrograde Warm Blood Cardioplegia: Myocardial Benefit and Neurologic Threat. Ann Thorac Surg. 1994; 57:298–304.

7. Durandy YD. Is there a rationale for short cardioplegia re-dosing intervals? World J Cardiol. 2015; 26; 7(10): 658–664.

8. Bayandin N.L. Crystalloid hypothermic and blood normothermic cardioplegia. Clin Experiment Surg. Petrovsky J. 2020; 8 (1): 80–9. doi: 10.33029/2308-1198-2020-8-1-80-89 (in Russian).

9. Carvajal C, Goyal A, Tadi P. Cardioplegia. In: StatPearls. Treasure Island (FL): StatPearls Publishing; January 31, 2021. https://www.ncbi.nlm.nih.gov/books/NBK554463/

10. Roques F, Michel P, Goldstone AR, Nashef SA. The logistic EuroSCORE. Eur Heart J. 2003 May;24(9):882–3. doi:10.1016/S0195-668X(02)00799-6

11. Siregar S, Groenwold RH, de Mol BA, Speekenbrink RG, Versteegh MI, Brandon Bravo Bruinsma GJ, Bots ML, van der Graaf Y, van Herwerden LA. Evaluation of cardiac surgery mortality rates: 30-day mortality or longer follow-up? Eur J Cardiothorac Surg. 2013 Nov;44(5):875–83. doi: 10.1093/ejcts/ezt119.

12. Mazzeffi M., Zivot J., Buchman T., Halkos M. In-Hospital Mortality After Cardiac Surgery: Patient Characteristics, Timing, and Association With Postoperative Length of Intensive Care Unit and Hospital Stay. Ann Thorac Surg 2014;97:1220–6. doi.org/10.1016/j.athoracsur.2013.10.040

13. Manlhiot C, Rao V, Rubin B, Lee DS. Comparison of cardiac surgery mortality reports using administrative and clinical data sources: a prospective cohort study. CMAJ Open. 2018;6(3):E316–E321. doi:10.9778/cmajo.20180072

14. Ota T, Yerebakan H, Neely RC, et al. Short-term outcomes in adult cardiac surgery in the use of del Nido cardioplegia solution. Perfusion. 2016; 31(1):27–33.

15. Lichtenstein SV, Naylor CD, Feindel CM, et al. Intermittent Warm Blood Cardioplegia. Circulation. 2018; 92 (9):341–346.

16. Ghazy T, Allham O, Ouda A, et al. Is repeated administration of blood-cardioplegia really necessary? Interact Cardiovasc Thorac Surg. 2009; 8(5):517–21.

17. Molina GR, Correa JR, Rios EG, et al. Single dose cold blood cardioplegia in adult patients: rationale and technique description. Cardiothorac Vasc Sci. 2018, 2(1): 1–4.

18. Sorabella RA, Akashi H, Yerebakan H, et al. Myocardial Protection Using Del Nido Cardioplegia Solution in Adult Reoperative Aortic Valve Surgery. J Card Surg. 2014; 29:445–449

